# Adapting, piloting, and evaluating a pediatric lead screening and risk-reduction intervention in Nairobi: A hybrid implementation-effectiveness trial protocol

**DOI:** 10.64898/2026.04.28.26351918

**Authors:** Ikenna Onoh, Cyrus Mugo, Anne Riederer, Elizabeth Maleche-Obimbo, Faridah Hussein Were, Christine Loftus, Ferdinand Mukumbang, Edith Lumumba, Barbra Richardson, Priscilla Wanini Edemba, Beatrice C Mutai, Catherine Karr, Sarah Benki-Nugent

## Abstract

**Background:** Childhood lead exposure is prevalent worldwide including low- and middle-income countries (LMICs). Structured screening and prevention programs to address pediatric lead exposure are largely absent in these settings. Adapted interventions are needed to close this implementation gap in an urban African context. This paper describes the protocol for the Lead Exposure Intervention Program (LEIP), which aims to adapt, pilot, and evaluate a pediatric lead exposure screening and risk-reduction protocol in Nairobi, Kenya.

**Methods:** LEIP is a multi-phase, hybrid type 3 implementation-effectiveness study. Phase 1 is a formative one-arm study leveraging an existing mother–child cohort and stakeholder-led tools adaptation to pilot a program comprising blood lead level (BLL) screening with a lead risk survey and tailored caregiver risk reduction messaging. Phase 2 is a randomized trial in public sector clinics. In this phase, approximately 1,500 children will be screened to identify 100 with elevated BLL (≥5 µg/dL) for enrollment, who will then be randomized 1:1 to receive either clinic-only risk-reduction messaging or the same clinic-based messaging plus a home visit for environmental assessment and additional tailored messaging. Follow-up at 3 and 9 months will assess caregiver recall of key messages and adoption of recommended exposure-reduction behaviors, as well as changes in child BLL. Phase 3 involves qualitative interviews with caregivers and key stakeholders to identify multi-level barriers and facilitators to intervention uptake. Quantitative and qualitative findings will be integrated to inform refinements for scale-up.

**Discussion:** This study represents a critical opportunity to develop and evaluate an adaptive, screening-based lead exposure intervention tailored to the urban LMIC context. By incorporating implementation science principles and stakeholder-driven design, LEIP is well-positioned to inform scalable national and regional approaches. The inclusion of both quantitative and qualitative components enhances the protocol’s ability to capture multilevel dynamics of uptake, fidelity, and sustainability, and generate actionable insights for future large-scale implementations.

**Trial Registration:** Registered on ClinicalTrials.gov (NCT07401251)

## Introduction

Lead poisoning in children has emerged as a critical global health issue, with mounting evidence that pediatric lead exposure is more prevalent than previously recognized. Recent estimates show that up to one in three children worldwide — approximately 800 million — have blood lead levels (BLLs) at or above 5 µg/dL [1], the current guidance level at which international agencies recommend intervention [2]. The burden of lead exposure is particularly significant in low- and middle-income countries (LMICs), where lead exposure controls and public health programs are often lacking [1]. Africa exemplifies this disparity: while children’s BLLs have declined dramatically in high-income countries [3], many African children today carry a heavy burden of lead [1,4–9]. Researchers estimate that childhood lead exposure in Africa results in the loss of around 98 million IQ points and economic costs of $134.7 billion annually (≈4% of African GDP) [10]. By contrast, the estimated annual economic loss due to lead exposure in the United States is about $50–55 billion [10]. Kenya, like many African countries, bears a significant burden of lead poisoning. Approximately 2.8 million Kenyan children have BLLs ≥5 µg/dL (with an estimated 326,000 above 10 µg/dL) according to UNICEF/Pure Earth estimates [1].

Lead is a potent neurotoxicant, and no lower threshold of safe exposure has been identified. Even low-level lead exposure is associated with irreversible cognitive and behavioral deficits [11–17]. Research demonstrates that the risk of adverse cognitive and behavioral outcomes increases with increasing BLL, even at very low levels [14–17]. Studies have linked early-life exposure to increased risks of attention disorders, reduced academic performance, and lifelong social and health burdens [12]. In LMICs, where children often face cumulative adversities — such as undernutrition, infectious disease, and limited educational resources — the additive impact of lead-related neurotoxicity has the potential to exacerbate inequities in child development [18].

In Sub-Saharan African cities, children’s lead exposure arises from a complex mixture of environmental, industrial, and household sources. Informal recycling of used lead-acid batteries (ULABs) and e-waste processing are contributors, releasing fine lead particulates and dust into communities [19,20]. Legacy contamination from vehicular emissions (before the phase-out of leaded gasoline) and industrial activities add to the cumulative load, especially in densely populated informal settlements [20]. Within households, children may ingest lead through peeling paint, contaminated soil and house dust, lead-glazed ceramics, artisanal cookware, and water pipes with lead solder [21]. Market surveys across 25 LMICs that analyzed 5007 individual products across 11 product types showed that over 50% of metal foodware and nearly 45% of ceramic items tested exceeded safe lead thresholds [21]. Spices, cosmetics, and traditional medicines containing lead are also potential sources of pediatric exposure in urban African settings [20].

Because of these combined exposures, urban African children may have higher BLLs than their rural counterparts [22–24]. Yet data on pediatric blood lead levels in African cities have until recently been sparse. The studies that have been conducted present a concerning picture. For instance, in Nairobi’s Kibera settlement, a 2009 survey of 387 children found that 7% of young children had BLLs ≥10 µg/dL (the prior threshold for “elevated” blood lead) [25]. Another study in 2014, in a lead-smelter-impacted community in Mombasa, Kenya, reported that 31% from a sample of 65 local children had BLLs ≥10 µg/dL [26]. By comparison, <1% of U.S. children exceed 10 µg/dL today [3].

Despite these indicators, structured programs to address pediatric lead exposure are virtually absent in LMICs. High-income countries (HICs), by contrast, have established surveillance, screening, prevention, and remediation programs for pediatric lead exposure. Through policies banning leaded gasoline and lead-containing paint, wide-scale housing abatement programs, and routine pediatric BLL screening, countries such as the United States have achieved dramatic declines in childhood lead poisoning — from 88% prevalence of BLL ≥10 µg/dL in the 1970s to under 1% in the 2000s [3]. In contrast, there are few national childhood lead screening or surveillance programs, minimal environmental monitoring, and limited capacity for household and community hazard remediation in most LMICs [27]. Policymakers in LMICs often underestimate the burden of lead poisoning because of scarce data and competing priorities. The implementation gap is profound: whereas HICs treat lead prevention as a routine public health practice, LMICs largely treat lead exposure as an episodic concern [27].

The Lead Exposure Intervention Program (LEIP) is designed to help close this programmatic gap. LEIP is a formative, implementation science study to adapt, pilot, and evaluate a pediatric lead screening and risk-reduction intervention in Nairobi, Kenya. This is an excellent setting due to its mix of high lead risk urban environments and health infrastructure that can be leveraged for interventions. By integrating accessible blood lead testing, a locally adapted risk-assessment survey, environmental assessment, and tailored caregiver risk-reduction messaging into existing health system and community workflows, LEIP seeks to establish a context-appropriate, scalable model. A recent one-arm longitudinal pilot intervention in Abidjan among pregnant women, some of whom also had other children, demonstrated that tailored information on the presence of lead-based paint in the home can improve awareness and enhance the adoption of self-reported preventive behaviors [28]. Rather than imposing imported interventions, LEIP emphasizes iterative adaptation and community engagement, aiming to refine intervention components based on real-world acceptability, feasibility, and potential integration. The long-term goal is to lay a foundation for sustainable implementation in Kenya and, by extension, in other African urban settings with similar exposure challenges.

In the spectrum of lead control strategies (**Figure 1**), primary prevention—eliminating lead hazards before exposure—remains the priority [29]. However, in resource-constrained contexts with existing contamination and exposure, primary prevention often requires long-term policy shifts, infrastructure investment, and legislative action. LEIP adopts a secondary prevention approach: identifying children with elevated BLLs through screening tests and intervening to halt further accumulation and mitigate downstream impacts [29]. This includes offering guidance to parents on reducing exposure (e.g., cleaning practices, avoiding certain cookware, dietary interventions), and providing follow-up BLL rechecks and referrals for appropriate medical care. While secondary prevention does not replace primary prevention strategies, it offers immediate protective benefits to exposed children and can generate critical empirical data to inform primary prevention strategies. By pairing individual-level interventions with data-driven advocacy, LEIP aspires to be a bridge, protecting children now while generating evidence and momentum for broader lead elimination.

**Fig. 1:**
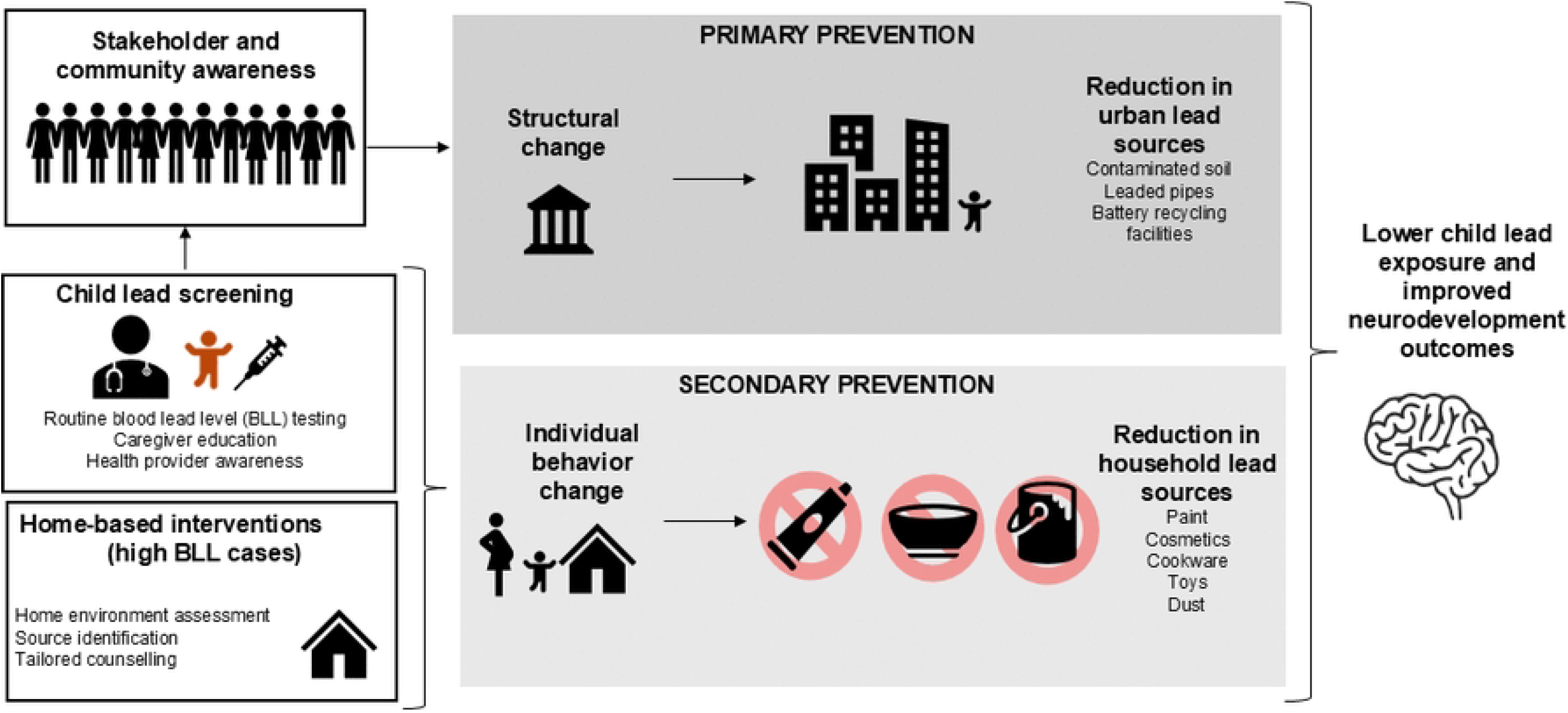
LEIP Study Conceptual Framework.

This formative protocol outlines how LEIP will pursue three integrated aims: (1) adapt, pilot and refine a prototype child blood lead screening, lead exposure risk survey, and messaging protocol; (2) evaluate uptake of tailored risk reduction messaging delivered in clinic only vs delivered with a home visit; and (3) evaluate individual, household, and structural barriers to uptake and sustained adoption, informing subsequent iterations and scale-up strategies. To our knowledge, this is the first such childhood lead exposure screening implementation science study in SSA. Insights gained will inform adaptation of larger-scale lead poisoning prevention programs in similar settings.

## Materials and methods

### Study design and setting

The LEIP study is structured as a multi-phase, hybrid type 3 implementation-effectiveness trial [30]. Three phases will be conducted: a formative one-arm pilot, a randomized trial of two implementation strategies, and mixed methods process evaluation. The process evaluation engages parents involved in the trial and key stakeholders (health workers and policymakers) in qualitative assessments.

The research is set in Nairobi, Kenya, based within public sector health facilities that provide routine maternal and child health services. The one-arm pilot leverages mother-child pairs participating in the Air Pollution Exposures in Early Life and Brain Development in Children (ABC) Study [31], a longitudinal pregnancy cohort study based at the Dandora II Health Centre, which is situated in the Dandora neighborhood and near the largest municipal dumpsite for Nairobi. The LEIP randomized trial will be based in Dandora II and additional publicly-accessible health facilities in Nairobi with demographically similar clientele. These sites will be selected to prioritize sites with a range of potential lead sources and/or where the study team has established study sites. Intervention delivery staff will include trained clinic and field personnel who have completed study-specific training in blood lead testing, risk reduction messaging, household observation procedures, and research ethics.

### Study governance and oversight

Study oversight will be provided by the investigative team in coordination with the LEIP Stakeholder Advisory Group, which includes representatives with expertise in pediatrics, environmental health, implementation science, public health practice, and policy in Kenya. This group will provide technical input on contextual adaptation, implementation considerations, and interpretation of findings. No separate endpoint adjudication committee is planned.

### Study status and timeline

LEIP is currently in its start-up phase. The study has not completed participant recruitment or primary data collection, and no study results are yet available. To date, study activities have been limited to protocol development, tool and messaging refinement, stakeholder engagement, staff preparation, and human subjects approvals. The overall study period is May 1, 2025, to April 30, 2030. Recruitment for Phase 1 is anticipated to be completed within one year of field implementation start, and recruitment for Phases 2 and 3 is anticipated to be completed within a subsequent one-year period, consistent with the approved study timeline. Final study data collection, including follow-up activities, is expected to be completed before the end of the grant period, and study results are expected thereafter.

### Phase 1: LEIP Tool Adaptation and Pilot Survey on Lead Exposure

#### Design, participants, and outcomes

The pilot consists of a one-arm study to adapt and evaluate a blood lead screening protocol which includes a lead risk survey on potential household lead exposure sources and complementary tailored messaging on lead risk reduction for urban Kenyan parents **(Figure 2)**. All parent-child pairs who were enrolled in the ABC study will be eligible for enrollment. The ABC children are anticipated to be between 36 and 48 months of age at the time of entry into the LEIP pilot. Up to 350 parent-child pairs will be enrolled. Primary outcomes of interest are understandability and acceptability of the blood lead testing, the risk survey, lead risk reduction message protocol, and accuracy of the survey.

**Fig. 2:**
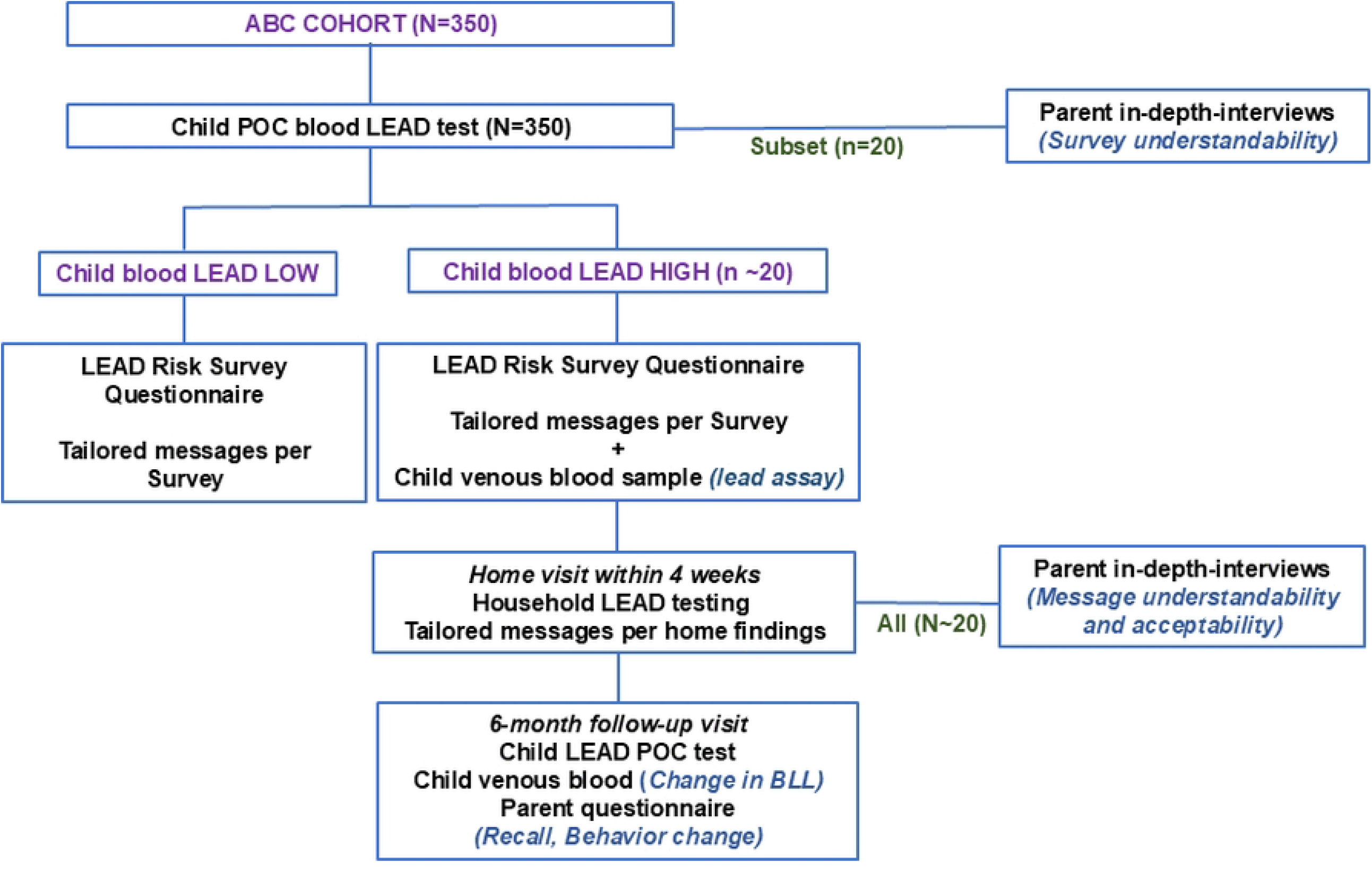
LEIP Study Phase 1 enrolment, follow-up and procedures. **Note:** Bold capital, **GROUP;** bold lowercase, **Procedure;** bold italics, ***Outcome*;** ABC, Early Exposure to Air Pollution and Brain Development in Children; POC, point-of-care; BLL, blood lead level

#### Tool adaptation

The survey questionnaire and messaging are prototypes, based on U.S.-based childhood lead poisoning prevention programs and globally relevant protocols [32–35], documented local sources [25,36–43], and expert input of local and global stakeholders.

#### Pilot Procedures

##### Lead risk survey and tailored risk reduction messaging

Parent-child pairs (most often mothers) will be offered point-of-care (POC) child blood lead screening (described below) and will undergo a brief structured lead risk survey **(Case Report Form in S2 Appendix)** to ascertain potential lead exposure sources in the household. Based on their individual responses to this survey, all participants will receive both standard and additional risk reduction messaging, tailored to their specific risk profile. Results from blood lead screening will be provided immediately.

##### Blood lead level screening and specimen collection for children identified to have high BLL

Using standard protocols [44], POC blood lead screening will be done using a capillary (fingerstick) sample and LeadCare® II assays and devices (Meridian Bioscience, Cincinnati, USA). For children whose initial BLL is high (≥5 µg/dL, the current WHO reference level of concern), an immediate confirmatory POC blood lead screening will be done using a second capillary sample. For children with confirmed high BLL, a venous blood specimen will be collected for real-time anemia testing and archived for future highly sensitive assays for lead. Separate informed consent will be obtained for long-term storage and future use of biological specimens. Parents will be informed of the intended use of archived specimens, storage procedures, and their right to decline specimen storage without affecting participation in the main study.

##### Home visits

Parents of children with high BLL will be offered a home visit within 4 weeks for a detailed household lead assessment of potential household lead exposure sources. At this time, individually tailored messaging will also be provided per the home findings. A trained field worker will administer a home observational checklist in order to systematically observe potential household lead hazards in and in proximity to the home (i.e., compound), such as peeling and chipping paint, artisanal cookware, bare soil play areas near a roadway, metal or painted toys, cosmetics such as kohl, or proximal industrial activities (formal or informal) that may include lead emissions. Following manufacturer instructions and standard protocols [21,45], a portable X-ray fluorescence device (pXRF) will be used to test suspect items and surfaces, such as soil in play areas, cookware, or locally-made non-branded cooking spices and traditional medicinal powders, *in situ* for lead content. Observations will be used to evaluate consistency and agreement with parent self-reported information ascertained at the enrollment visit and used to refine the lead risk survey for accuracy.

##### Laboratory procedures for venous blood specimens

Venous blood samples will be used for two tests: i) Blood hemoglobin levels within 48-72 hours (∼ 1 milliliter) and ii) Stored for batched blood lead testing using the traditional “gold-standard” assay Inductively Coupled Plasma Mass Spectrometry (ICP-MS) in the laboratory at the Department of Chemistry in the University of Nairobi using standard protocols [46]. This shall provide a reference test to complement the findings from each child’s POC LeadCare ® II assay.

##### Evaluation of lead risk survey questionnaire and risk reduction message understandability and acceptability

In-depth exit interviews will be conducted among two subsets of parents from the pilot. These subsets will include selected parents following the lead risk survey and all home visit participants. These in-depth interviews will be conducted within 1-2 days after the lead risk survey (purposively selected subset, N = 20) and home visit (all home visit participants, N ∼20) and will employ cognitive interviewing techniques to assess the understanding and acceptability of the survey questions and the risk reduction messaging.

Interview guides are based on select constructs from Damschroder’s Consolidated Framework for Implementation Research (CFIR) [47] that are considered relevant to phase 1 outcomes **(Table 1)** along with the researcher’s experience. Sample questions for lead risk survey understandability are *“Were the questions easy to understand? Was it hard to know the answer to the questions? What was it about the questions that was not understandable / were they hard to know the answer? Did answering any questions make you feel annoyed / bored / uncomfortable / disrespected?”*

**Table 1:** Consolidated Framework for Implementation Research (CFIR) domains and select constructs for Phases 1 and 3 design of interview guides and analysis.

| Domain | Constructs | Phase |  |
| --- | --- | --- | --- |
|  |  | 1 | 3 |
| Intervention characteristics | Intervention source |  | x |
|  | Evidence strength & quality |  | x |
|  | Adaptability |  | x |
|  | Relative advantage |  | x |
|  | Trialability |  |  |
|  | Complexity |  | x |
|  | Design quality & packaging | x | x |
|  | Cost | x | x |
| Outer setting | Patient needs & resources |  | x |
|  | Cosmopolitanism |  |  |
|  | Peer pressure |  |  |
|  | External policy & incentives |  | x |
|  | Community characteristics | x | x |
| Inner setting | Structural characteristics |  | x |
|  | Networks & communications |  | x |
|  | Culture |  | x |
|  | Implementation climate |  | x |
|  | Readiness for implementation |  | x |
| Characteristics of individuals | Knowledge & beliefs about intervention | x | x |
|  | Self-efficacy | x | x |
|  | Individual stage of change | x | x |
|  | Individual identification with organization |  |  |
|  | Other personal attributes | x | x |
| Process | Planning |  | x |
|  | Engaging |  | x |
|  | Executing |  | x |
|  | Reflecting & evaluating |  | x |
|  | Resource source |  | x |

The exit interviews for the home visit participants will gather information on the understandability and acceptability of risk reduction messages, and explore participant anticipated facilitators and barriers to the uptake of recommended exposure reduction measures. In addition to using the interview guides, we will also ask survey questions on acceptability from the Acceptability of Intervention Measure (AIM), a brief validated scale. The AIM scale is easy to administer and exhibits strong psychometric properties [48]. Sample questions will include *“Are there any situations or things that will make it hard to make these changes? What kind of support would you need to help you make these changes? Are there things that you think may help?”*

##### Six-month follow-up visit: Recheck of child BLL and assessment of parent knowledge and behavior change

At 6 months post-enrollment, a BLL re-check will be done using the same procedure as for the initial test, and a venous blood draw will be performed. A knowledge, attitudes, and practice questionnaire will be administered. This will be used to assess the caregiver’s recall of the earlier lead exposure messaging, uptake of recommended reduction measures, and individual determinants of uptake. Questions will be based on the Risk Attitude Norm Ability Self-Regulation (RANAS) framework [49] **(Table 2)**, which has been used to develop interventions related to environmental health behaviors and in diverse LMIC settings, including in SSA [50–52], to assess psychosocial determinants of health behaviors.

**Table 2:** Risks, Attitudes, Norms, Abilities and Self-regulation (RANAS) Framework, Factors, Subdomains, Sample questions and topics relevant for lead testing and reduction of exposure to lead.

| Factor | Subdomains | Sample questions or topics |
| --- | --- | --- |
| **Risk** | Factual knowledge, perceived vulnerability, perceived severity | Mother’s knowledge about lead health concerns and concerns related to child’s exposure to lead |
| **Attitude** | Beliefs about costs and benefits, feelings | Feelings about usefulness of having child receive blood lead test and making changes if it is high |
| **Norm** | Other’s behavior, dis(approval), personal importance | Testing is a good thing to do for my baby; Other people should have their children tested |
| **Ability** | How-to-do knowledge, confidence in performance | I know some ways to protect my child from lead; I am able to do things to reduce my child’s exposure to lead |
| **Self-regulation** | Action planning, action control, barrier planning, remembering, commitment | I plan to reduce my child’s exposure by...<br>I have reduced my child’s exposure by...<br>Challenges for me are .... but I can manage that by... |

#### Analysis

Primary outcomes are survey understandability and acceptability, agreement between in-clinic administered survey responses and at-home observational checklist findings and understandability and acceptability of risk reduction messages. For the qualitative data, a hybrid inductive and deductive thematic analysis approach based on *a priori* selected CFIR constructs will be conducted.

For the quantitative data, we will explore the proportion of mothers (self-report on in-clinic surveys) and assessed homes with identified actionable targets, e.g., suspect household products, including cosmetics, chipped paint, potential occupational exposure, or other lead containing items observed using pXRF. We will describe the number and types of actionable targets identified for each participant. This data will enhance our adapted survey and messaging protocols for the randomized trial. We will summarize descriptive statistics for standardized questionnaires on recall of identified risk factors, behavior change (e.g., proportions with recall of all or most factors vs. proportions with recall of some or no factors), and attitude. We will select questions with higher variability, and we will refine our questionnaires as appropriate.

### Phase 2: Randomized Trial of Messaging Strategies

#### Design, intervention, and outcomes

Phase 2 will involve a randomized trial comparing two implementation strategies for the lead exposure risk reduction messaging. The trial will follow the checklist and guidelines described in the SPIRIT guidelines for the content of clinical trial protocols (**Figure 3** and Supporting Information in **S1 Checklist**). This study aligns with the hybrid type 3 design as we evaluate two implementation strategies concurrently while gathering information on clinical outcomes [30]. Primary outcomes will be uptake of BLL recheck visits (retention) and recall and uptake of messages. The exploratory outcome will be change in child BLL. **(Figure 4)**

**Fig. 3:**
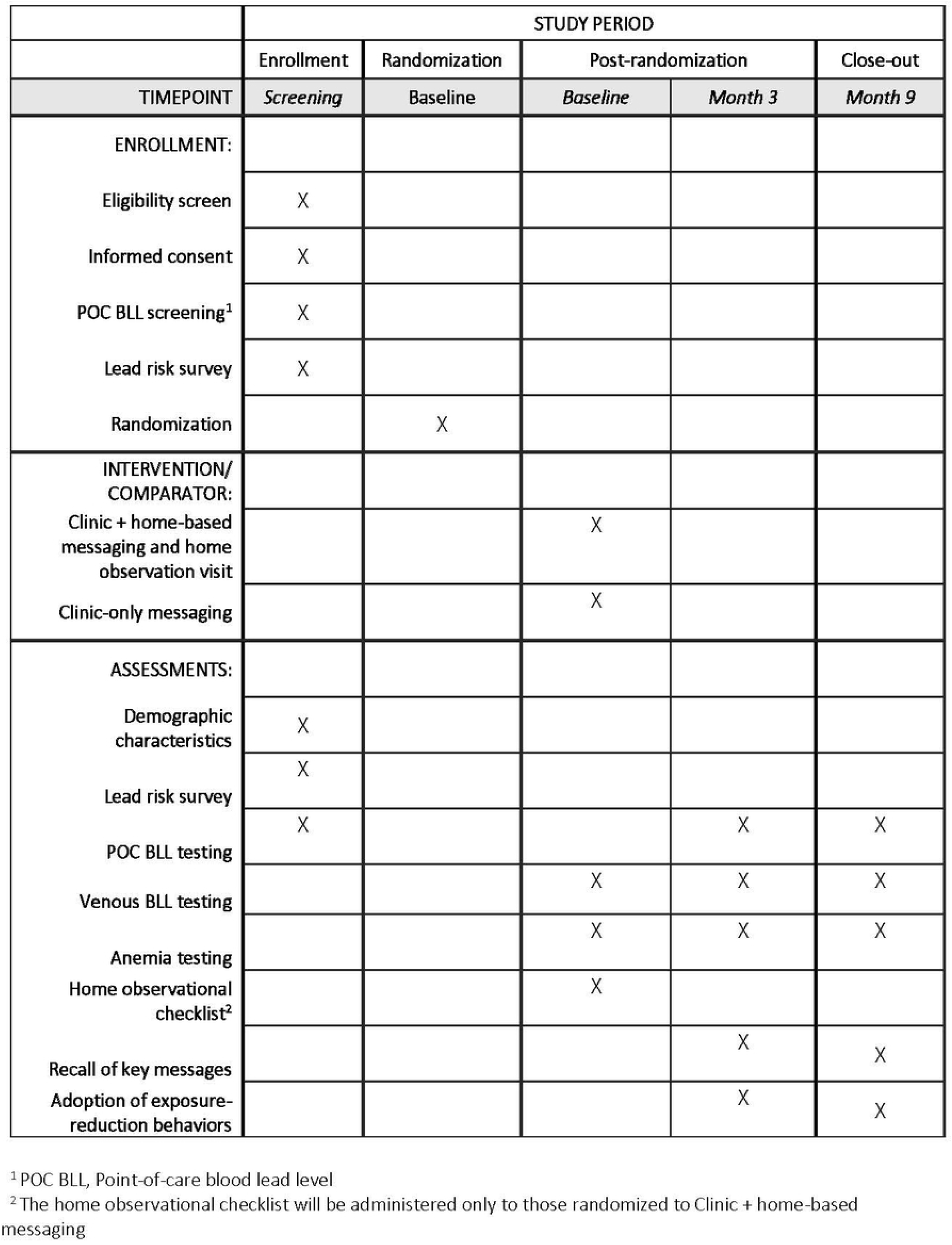
Schedule of enrollment, intervention, and assessments for the LEIP trial (Phase 2) as outlined in the SPIRIT recommendations.

**Fig. 4:**
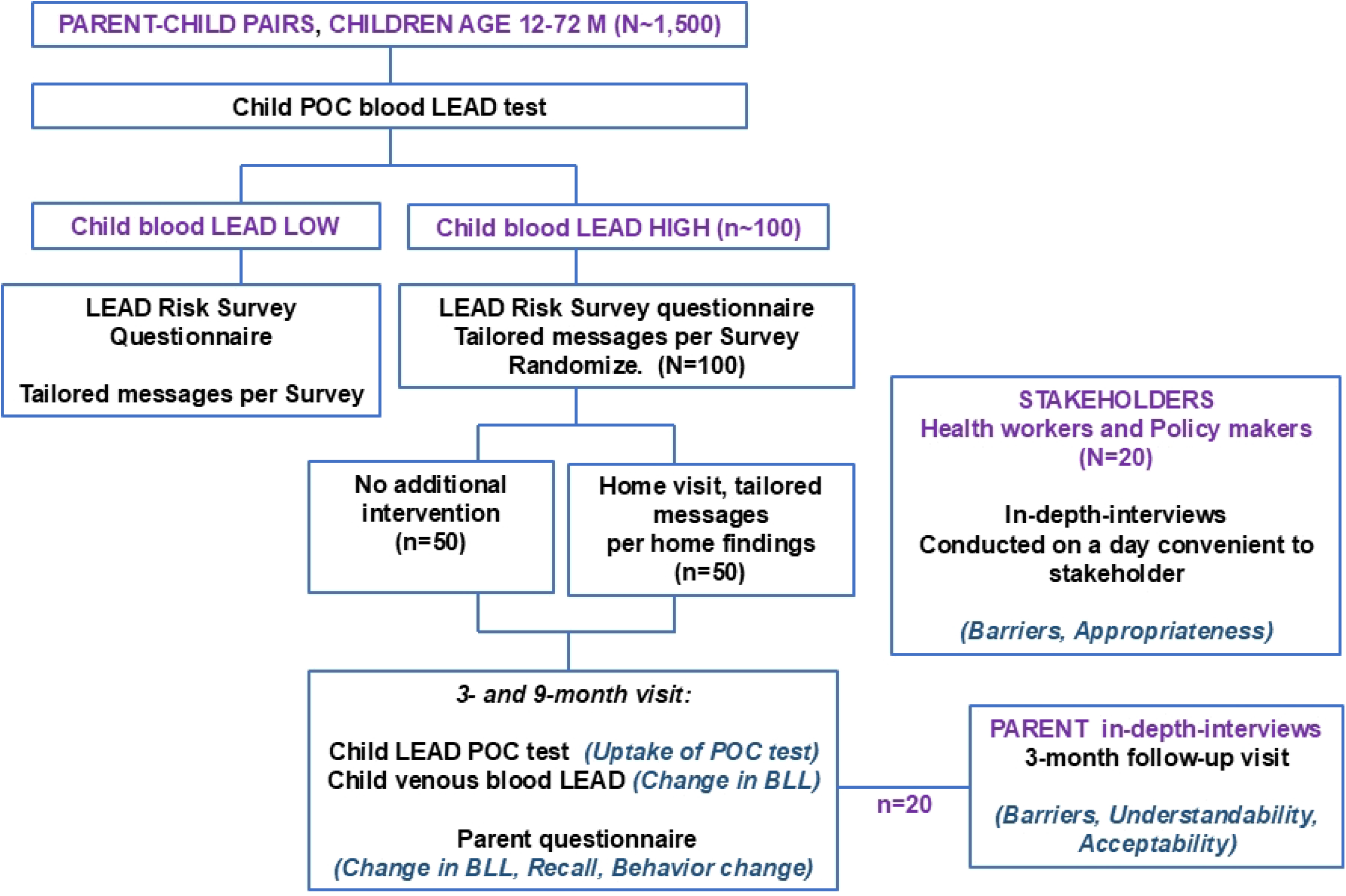
Phases 2 and 3 Schematic. **Note:** Bold capital, **GROUP;** bold lowercase, **Procedure;** bold italics, ***Outcome*;** POC, point-of-care; BLL, blood lead level

#### Procedures

##### Screening and enrollment

Key eligibility criteria for enrollment will be children aged 12-60 months and parents aged ≥18 years, and plan to reside within the study catchment area for at least 9 months (to allow for home visits and BLL rechecks). Parent-child dyads attending routine well-childcare in public sector clinics will be told about the study, screened for eligibility, and those consenting to participate shall be enrolled consecutively up to a maximum sample size of 1500 parent-child pairs. A POC blood lead test shall be done for each child until a target sample size for randomization of 100 children with high BLL is reached.

##### Procedures at enrollment

Following a brief questionnaire to ascertain demographic characteristics, all enrolled children will undergo POC BLL screening (described above), and immediate communication of the result. The standardized lead risk survey and risk reduction messaging, which will be honed based on the pilot, will be administered. Children with confirmed high BLL will have a venous blood sample collected for anemia testing and specimen archival for future assays (described above).

##### Randomization

Children with confirmed high BLL will be randomly assigned in a 1:1 ratio to one of two messaging strategies (N=50 per arm). Randomization will be within two sub-groups stratified by child baseline BLL, 5 to 10 ug/dL and >10 ug/dL. The random allocation sequence will be generated by the study biostatistician using a computer-generated randomization schedule. Allocation will be concealed from members of the study staff until assignment using sequentially numbered opaque sealed envelopes. Because of the nature of the intervention, participants and intervention delivery staff cannot be blinded to trial arm assignment. Arm 1 will involve clinic-only messaging. Families randomized to this arm will receive only the in-clinic risk reduction messaging after the child’s BLL screen, tailored based on the lead risk survey responses. Arm 2 will involve clinic + home-based messaging. Families in this arm will receive the same initial messaging at the clinic, *plus* a home observation visit within 4 weeks. The home visit protocol will include an observational checklist to identify and point out any lead sources and reinforce the risk messages in the context of the family’s actual home environment. Any hazards identified will be addressed with tailored guidance, and caregivers will be helped to problem-solve ways to implement recommendations. The clinic-only messaging arm was selected as the comparator because it represents the most feasible minimum implementation package within routine public-sector child health services. The clinic plus home-visit arm tests whether contextualized environmental assessment and reinforced tailored messaging improve uptake beyond what can be achieved through clinic-messaging alone.

##### Administration of concomitant care

Participants will continue to receive routine pediatric and maternal-child health services throughout the study. No standard clinical care will be withheld. Additional non-study services or counseling sought by families will not be prohibited and will be documented when relevant to interpretation of outcomes.

##### Follow-up visits: Recheck of BLL and assessment of parent knowledge and behavior change

All 100 trial participants will be scheduled for 3- and 9-month follow-up visits for POC BLL rechecks, and a venous blood draw for real-time anemia testing and specimen storage. A standardized questionnaire will ascertain: (1) parent recall of lead sources and hazards identified for their child; (2) the specific actions or changes the parent has carried out since the messaging was given; and (3) individual determinants related to these behaviors using the RANAS framework.

##### Safety and intervention modification

Participants may discontinue study participation at any time at their own request or at the request of their parent or guardian, without penalty. Study procedures may also be modified or discontinued if a child has clinical findings that warrant urgent referral, if repeated blood sampling is not feasible, or if a home visit is judged unsafe or unacceptable.

##### Strategies to improve and monitor adherence

To promote adherence to assigned intervention procedures and follow-up visits, the study team will use appointment reminders, flexible scheduling, caregiver contact verification, and reimbursement for transport and time. Intervention delivery and follow-up completion will be documented using standardized study forms, including completion of messaging, home visits, and scheduled BLL rechecks. If participants discontinue intervention activities but do not withdraw consent, follow-up outcome data will continue to be collected whenever feasible.

##### Data management

All study data will be entered into a secure, password-protected database with role-based access controls. Standardized coding procedures, range and logic checks, routine review of missingness, and periodic quality-control checks will be used to promote data quality. Identifiable information will be stored separately from analytic data and retained only for as long as necessary under institutional and ethical requirements.

#### Sample size and analysis

Based on a prevalence of 8.2% of high BLL from preliminary data in children enrolled in ABC, we will enroll and screen 1,500 children to attain 100 trial participants. The trial’s sample size (N=100) was determined by practical considerations and precision for estimating uptake and recall. With 90 participants expected to complete the 9-month follow-up (assuming 10% attrition) and observed

≥67% of parents in an arm implementing recommendations, we will have ≥ 80% power to exclude a recall or uptake of < 50%. Similarly, the study can detect a meaningful difference between a moderate uptake rate (e.g. ∼50%) and a poor uptake (<25%). The primary outcomes of recall and behavior uptake at 3 and 9 months will be reported as proportions with 95% CIs in each arm and stratified by initial BLL category. We hypothesize generally high uptake in both arms, with potentially higher rates among those with BLL >10 µg/dL. Between-arm comparisons will be examined using χ^2^ or Fisher’s exact tests for proportions, acknowledging that we will have limited ability to exclude small differences.

Primary quantitative analyses will include all randomized participants with available follow-up data according to their assigned study arm. Additional exploratory analyses may be restricted to participants who complete key intervention components, as specified *a priori*. Missing data are anticipated to be minimal. Primary analyses will therefore use a complete-case approach, with participants excluded from a given model only if data required for that analysis are missing. The extent and pattern of missingness will be summarized, and characteristics of included and excluded participants will be compared descriptively to assess the potential for bias. For descriptive analyses, available data will be used and varying denominators will be reported where applicable.

We consider BLL change to be a distal indicator of effectiveness. In a prior Kenyan study, standard deviations (SDs) were 15.5 ug/dL for maternal and 1.9 ug/dL for cord BLL [46]. We anticipate that the SDs for child BLL will be lower than for adult women but higher than for cord samples. Assuming 10% attrition, alpha = 0.05, a two-sided test, ≥ 80% power, and a paired t-test with a correlation of 0.8, we will be able to detect a difference in −0.37 ug/dL between arms, for SD = 2 and a difference of −2.83 ug/dL between arms, for SD = 15. We acknowledge this study may not have adequate power nor follow-up time to detect a significant reduction in child BLL, given that only a subset of parents may have the ability to identify modifiable risk reduction behavior.

We will explore associations between the hypothesized individual determinants (e.g., knowledge acquisition, empowerment and self-efficacy, as well as male partner involvement and caregiver sex) and the uptake of behaviors. Specific indicators will include perceived vulnerability, perceived risk, attitude regarding cost of lead exposure reduction, and perceived ability to take up messages. We hypothesize that caregivers with uptake of risk reduction behavior will have higher perceived vulnerability and risk, and greater perceived ability to take up messages compared with caregivers with lower uptake. We will use Wilcoxon rank-sum tests and log binomial regression or modified Poisson regression with robust errors, as appropriate to explore the relation between Risk, Attitude, and Ability scores and uptake of risk reduction messages. These analyses will help identify which psychosocial factors are most strongly associated with successful behavior change, informing the design of future interventions.

The full study protocol and supporting instruments are provided as Supporting Information (**S3 Appendix. Study protocol**). No separate statistical analysis plan has been finalized at this stage; planned analyses are described in the present manuscript. If a separate statistical analysis plan is developed, it will be made available through the trial registry or journal supplementary materials.

### Phase 3: Qualitative Evaluation of Barriers and Facilitators to the Uptake of Lead-Reduction Messaging

We will use qualitative methods to understand the barriers and facilitators affecting message uptake at both individual and structural levels, to inform policy solutions. Following the 3-month follow-up, we will include parents of children with a range of BLL levels (e.g. 5–10 vs >10 µg/dL) To guide the inquiry, we will draw on CFIR constructs and the RANAS framework. Interview guides will explore knowledge acquisition, empowerment and self-efficacy, family dynamics and the socioecological context of these factors, to enable mapping of these factors according to structural, community, household, individual, and habitual levels [52].

Across the entire study period and with flexible scheduling at their convenience, we will conduct key informant interviews with stakeholders to capture broader systemic barriers. We will recruit individuals identified via our stakeholder advisory network, including frontline health providers (e.g., nurses, community health workers) involved in LEIP implementation, local public health and environmental officials (e.g. representatives from the Ministry of Health, or the Nairobi County), and community leaders or NGO representatives concerned with environmental health. Interview guides will be developed using CFIR constructs **(Table 2)** selected for the purpose of evaluating determinants of risk reduction practices.

### Monitoring and safety oversight

No independent data monitoring committee is planned because this is a low-risk behavioral and implementation study with minimal anticipated adverse events. Safety oversight will instead be provided by the study investigators in coordination with institutional ethics oversight and predefined referral procedures for children with clinically important findings. No formal interim analyses or stopping guidelines are planned for this study. However, serious unanticipated concerns related to participant safety, data integrity, or feasibility will be reviewed by the investigative team and relevant oversight bodies. Trial conduct will be monitored internally through regular team meetings, review of recruitment and follow-up performance, verification of key study procedures, and routine data quality checks. Any protocol deviations or implementation challenges identified during monitoring will be documented and addressed through retraining or procedural clarification as needed.

### Ethical considerations

This study has been approved by the Kenyatta National Hospital-University of Nairobi Ethics and Research Committee in Kenya and by the University of Washington Institutional Review Board (IRB) in the United States. We will also obtain clearance from relevant county and health facility authorities in Nairobi before implementation. The trial is registered at ClinicalTrials.gov (NCT07401251). Important protocol modifications will be submitted for review and approval by the relevant ethics committees before implementation, except where immediate action is required to protect participant safety. Approved amendments will be communicated to investigators, trial registries, and other relevant parties as appropriate. Potential harms related to study participation are expected to be minimal and include transient discomfort or bruising from blood collection, caregiver distress related to elevated blood lead findings, and possible social or household tension related to discussion of environmental hazards. These events will be documented by the study team and managed through counseling, referral, clinical follow-up, or modification of study procedures as appropriate. There is no compensation available for study-related harm. As direct benefit to the participants, children identified to have high BLL, anemia, or other clinically relevant findings during study procedures will be evaluated by study clinicians, and where clinically indicated be referred to the referral hospital for further management. In addition, tailored health education on potential sources of lead exposure identified from the home evaluation and parent interviews shall be provided to empower the parents to reduce lead exposure risk for their child. Personal information about potential and enrolled participants will be collected only as necessary for study conduct and follow-up. Any dissemination of results will use de-identified, aggregate data. Study results will be disseminated to the local community and stakeholders through report-back meetings and policy briefs, and globally through conference presentations and open-access publications.

## Discussion

This protocol outlines the methods for a formative, multi-phase implementation science study to adapt, pilot, and evaluate a pediatric lead screening and risk-reduction intervention in Nairobi, Kenya. We will adapt and contextualize elements from established childhood lead poisoning prevention programs in high-income countries (HICs), notably provision of accessible blood lead level testing, identification of potential sources contributing to higher BLLs, corresponding risk reduction messaging, and BLL follow up if levels are above guidelines.

Adapting high-income countries (HIC) lead screening and prevention programs to an urban African context leverages well-developed strategies but must contend with local constraints and previous barriers. For example, training local staff in lead testing and risk messaging increases sustainability [1], as it builds long-term workforce capacity in environmental health that is currently underdeveloped. In addition, laboratory infrastructure in Kenya is limited: few laboratories can perform blood lead assays. We overcome this by using portable POC analyzers (LeadCare® II), used widely in HIC settings for screening programs and previously employed in research studies in LMICs [53–58]. For elevated samples, confirmatory testing using a second POC test provides a practical approach for settings without ready access to more sophisticated laboratory methods.

Evaluating a lead screening program in the context of routine child health visits emulates screening programs in the USA. Our risk survey will be derived from prior lead exposure risk surveys used in LMIC investigations [32] and existing evidence of risk factors in LMICs. Our prototypes will be co-developed with local experts, enhancing cultural relevance. Furthermore, our design incorporates cognitive testing of survey items and validates responses via home visits. In Nairobi, these approaches set the stage for future programs integrated into existing services.

Our trial design seeks to inform policymakers of the minimum implementable package appropriate for this context. An intervention package including a home visit may be most effective, however an in-clinic risk survey with risk reduction messaging alone may be the most feasible and warrants evaluation. Our randomized design will allow us to evaluate both models in similar groups of parent-child pairs to inform policymakers on the minimum implementable package appropriate for this context. Our mixed-methods exploration of barriers and facilitators (Phase 3) will inform the next steps beyond this implementation study. Qualitative interviews with caregivers and health workers may reveal socio-economic and cultural barriers to risk reduction that quantitative data alone cannot capture. With systematic training of interviewers and in-depth interview guides, we will gain robust insights that can be triangulated with quantitative outcomes to validate results.

Generating local data and engaging policy makers will contribute to complementary needs for future policy scale efforts, including primary prevention activities. For example, involving government officials will ensure our work is policy relevant. Interim findings will be shared with decision-makers, framing results in terms of child health and development, and collaborating with awareness campaigns. In the short term, we will measure immediate outcomes as proxies for longer-term impact, recognizing that structural changes will require sustained effort beyond this study.

In summary, we leverage the strengths of an established research collaborative linking HIC experts in pediatric environmental health, lead screening, and implementation science with Nairobi-based pediatric health and lead exposure specialists to provide critical evidence needed for meaningful and sustainable implementation of lead screening in an urban SSA context.

## Supporting Information

**S1 Checklist. SPIRIT 2025 checklist**

**S2 Appendix. Case report form**

**S3 Appendix. Study protocol**

## Author Contributions

**Conceptualization:** Ikenna Onoh, Cyrus Mugo, Anne Riederer, Faridah Hussein Were, Christine Loftus, Ferdinand Mukumbang, Elizabeth Maleche-Obimbo, Catherine Karr, Sarah Benki-Nugent

**Funding Acquisition:** Elizabeth Maleche-Obimbo, Catherine Karr, Sarah Benki-Nugent

**Methodology:** Cyrus Mugo, Anne Riederer, Faridah Hussein Were, Christine Loftus, Ferdinand Mukumbang, Edith Lumumba, Barbra Richardson, Elizabeth Maleche-Obimbo, Catherine Karr, Sarah Benki-Nugent

**Project Administration:** Priscilla Wanini Edemba, Beatrice C Mutai, Elizabeth Maleche-Obimbo, Catherine Karr, Sarah Benki-Nugent

**Supervision:** Priscilla Wanini Edemba, Beatrice C Mutai, Elizabeth Maleche-Obimbo, Catherine Karr, Sarah Benki-Nugent

**Visualization:** Ikenna Onoh, Cyrus Mugo, Christine Loftus, Ferdinand Mukumbang, Elizabeth Maleche-Obimbo, Catherine Karr, Sarah Benki-Nugent

**Writing – Original Draft Preparation:** Ikenna Onoh, Catherine Karr, Sarah Benki-Nugent

**Writing – Review & Editing:** Ikenna Onoh, Cyrus Mugo, Anne Riederer, Faridah Hussein Were, Christine Loftus, Ferdinand Mukumbang, Edith Lumumba, Barbra Richardson, Priscilla Wanini Edemba, Beatrice C Mutai, Elizabeth Maleche-Obimbo, Catherine Karr, Sarah Benki-Nugent

## Data availability statement

No datasets were analyzed or reported in the current manuscript. De-identified participant data, data dictionaries, and analytic code will be made available after publication upon reasonable request and in accordance with applicable ethical, legal, and institutional requirements. Requests will be reviewed by the study investigators and relevant institutional oversight bodies.

